# Increased Spatiotemporal Variability after Stroke is not Just the Outcome of Walking Velocity

**DOI:** 10.1101/2023.02.03.23285403

**Authors:** Yogev Koren, Oren Barzel, Lior Shmuelof, Shirley Handelzalts

**Affiliations:** Department of Physical Therapy, Faculty of Health Sciences, Ben-Gurion University of the Negev, Beer-Sheva, Israel; The Translational Neurorehabilitation Laboratory, Adi-Negev Nahalat Eran Rehabilitation Center, Ofakim, Israel; Sheba Medical Center, Ramat Gan, Israel; Adi-Negev Rehabilitation Center, Nahalat Eran, Israel; Sackler Faculty of Medicine, Tel Aviv University, Tel Aviv, Israel; Ono Academic College, Kiryat Ono, Israel; Department of Cognitive and Brain Sciences, Ben-Gurion University of the Negev, Beer Sheva, Israel

**Keywords:** Walking, coordination, LEMOCOT, motor control

## Abstract

**Background:** Increased spatiotemporal gait variability is considered a clinical biomarker of ageing and pathology, and a predictor of future falls. Nevertheless, it is unclear whether the increased spatiotemporal variability observed in persons with stroke (PwS) is directly related to the pathology or simply reflects their choice of walking velocity.

**Research question:** Does increased spatiotemporal gait variability directly relate to motor coordination deficits after stroke?

**Methods:** Forty PwS and 20 healthy adults participated in this cross-sectional study. Participants performed the lower-extremity motor coordination test (LEMOCOT) on an electronic mat equipped with force sensors. Then, participants walked for 120 s over a computerized treadmill at their comfortable walking velocity. For the LEMOCOT we used the traditional score of in-target touch count and computed the absolute and variable error around the targets. For gait variability, we extracted the standard deviation of step time, step length, step velocity, and step width. Using a data-based approach, we generated a model of the relationship between velocity and variability and tested the correlations of the velocity-controlled values with the outcome measures from the LEMOCOT.

**Results:** PwS demonstrated increased variability in step time, step length, and step velocity, as well as decreased variability in step width, in comparison to healthy adults, even after controlling for walking velocity. After controlling for walking velocity, we observed that for PwS the LEMOCOT score correlated with the variance in step time, and the variable error in the LEMOCOT correlated with the variance in step length, in step width, and in step velocity. No significant correlation with any of the velocity-controlled step parameters was found for the absolute error in the LEMOCOT.

**Significance:** Decreased performance in the LEMOCOT was associated with increased spatiotemporal variability in PwS, regardless of their walking velocity. Our results demonstrate the connection between lower-extremity coordination impairments and deficits in gait function.

## 1. Introduction

Falls are a common complication after a stroke, with significant physical and psychosocial consequences that contribute to decreased independence and poor quality of life [1,2]. In community-dwelling persons with stroke (PwS), walking is the most frequently mentioned activity at the time of a fall [2]. Identifying PwS at risk for falls is of significant clinical importance given the effort directed toward balance and gait training, as well as preventing falls, throughout the rehabilitation process.

Several indices to assess gait stability have been suggested and used in the literature (e.g., [3]). One of the most used indices is the variability in spatiotemporal parameters (e.g., step/stride time variability and step/stride length variability [3]). Generally, increased spatiotemporal variability (STV) is considered an indication of greater instability and reduced ability to attenuate small perturbations [3], or in other words, “less” is good and “more” is bad [4]. This perspective is supported by a growing body of literature identifying gait variability as a clinical biomarker of ageing [5,6] and pathology [7,8] as well as a predictor of future falls [9,10].

Nevertheless, STV is sensitive to walking velocity in both healthy [11-14] and patient populations [15,16]. Given that slow walking is also a biomarker of pathology, it is unclear whether the increased gait variability observed in patient populations such as PwS [17,18] is directly related to the pathology or simply reflects their choice of walking velocity (i.e., indirectly related to the pathology).

Motor coordination may be defined as the ability to produce organized movement in both spatial and temporal domains, in a context-dependent manner [19,20]. This ability is important during walking as the relationship between body segments needs to be adaptable to accommodate internal and external demands and to allow accurate foot placement and safe mobility [21,22]. Errors in step execution could disturb walking stability, especially when negotiating obstacles or cluttered walking paths.

Hence, we used the lower extremity motor coordination test (LEMOCOT) [23], a performance-based measure to assess lower extremity motor coordination. Previous reports have indicated that persons with neurological disorders, such as PwS, often demonstrate motor coordination deficits that may limit the performance of daily activities [24,25]. Further, the LEMOCOT has been reported to correlate with performance-based measures of walking velocity and capacity [23,26].

In the current study we aimed to test whether STV is directly related to motor coordination deficits following stroke and is not a consequence of decreased walking velocity. We used a data-based approach to model the velocity-variability relationship and then tested the correlation of these models’ residuals with motor coordination deficits as reflected by the LEMOCOT. We included a group of healthy adults for which the LEMOCOT score is unlikely to reflect deficits, but instead is more likely to reflect ability. In doing so, we hoped to provide further insights regarding the nature of STV. We hypothesized that STV would correlate with walking velocity in healthy adults and in PwS. Also, we predicted that PwS would demonstrate increased STV regardless of walking velocity and that after controlling for walking velocity, the STV parameters and the LEMOCOT score would correlate in PwS (i.e., increased gait variability would be associated with lower performance on the LEMOCOT) but not in healthy adults.

## 2. Methods

### Participants

Forty PwS and 20 healthy adults participated in this exploratory cross-sectional study. Participants in the stroke group were recruited during their hospitalization at Adi Negev Nahalat-Eran Rehabilitation Center in Israel. Inclusion criteria included first-ever unilateral ischemic or hemorrhagic stroke and ability to walk on a treadmill independently or under supervision for 2 min. Exclusion criteria included other musculoskeletal or neurological injuries, and pain that could interfere with the performance of the tasks. Healthy adults were recruited from among the hospital’s staff and visitors. All participants signed an informed-consent form. The study was approved by the regional ethical review board at Sheba Medical Center, Israel (approval number 6218-19-SMC).

### Study protocol

#### LEMOCOT

Participants sat on a chair and preformed the test barefoot, as described by Desrosiers et al. [23]. Participants were instructed to alternately touch the proximal and distal targets with their big toe as fast and as accurately as possible for 20 s. The proximal and distal targets were marked on an electronic mat equipped with force sensors (Zebris FDM-T Treadmill, *Zebris Medical GmbH*, Germany) as described previously by our group [27]. PwS performed the test first with their non-paretic leg, followed by their paretic leg, and healthy participants performed the test first with their dominant leg (i.e., the leg used to kick a ball), followed by their non-dominant leg. During the test a physical therapist counted the number of in-target touches that constitutes the test’s score.

### Gait

Participants walked for 120 s over a computerized treadmill system (Zebris FDM-T Treadmill, *Zebris Medical GmbH*, Germany) at their comfortable walking velocity (CWV). This velocity was determined based on the average of two trials of the overground 10-meter walk test. Force data was acquired at 120-Hz sampling frequency using the software provided by the manufacturer (Zebris FDM, version 1.18.40). A safety harness attached to an overhead support protected participants in the event of a loss of balance but did not support any body weight or restrict their movements during walking. Participants were instructed to use the handrail support as little as possible. A physical therapist stood on the side of the treadmill for supervision but did not provide any assistance. Participants wore their own sports shoes and PwS could use foot orthosis in case they needed ankle support.

### Data processing

Dedicated algorithms and MATLAB scripts (MathWorks Inc.) were developed and used to analyze force data obtained during the LEMOCOT and the gait assessment trials. For the LEMOCOT the script computes 1) the absolute error (AE), and 2) the variable error (VE) around each target [27]. For simplicity, we used the average across the two targets.

For gait assessment, the script identifies the heel strike as the first time point of force detection after the swing phase. *Step time* was determined as the time interval between the heel strike of one leg and the heel strike of the opposite leg. *Step length* was calculated as follows: First, we calculated the area under the curve of the velocity-by-time graph (actual instantaneous velocity of the treadmill). To this value we added the difference in the anterior-posterior positions of the two consecutive heel strikes. To calculate *step width* the algorithm computes the distance between the mediolateral position of the center of pressure (COP) during one heel strike and the COP position of the consecutive heel strike. *Step velocity* was calculated as 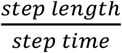. The standard deviation (SD) of all parameters was used as a measure of gait variability.

### Statistical analysis

Between-group comparisons of participants’ characteristics were performed using the Mann-Whitney U test. For the LEMOCOT analysis we included only the performance of the paretic leg in PwS and left leg in healthy participants. (It should be noted that for both groups this leg was tested second). To test whether the LEMOCOT score correlates with the 10-m walk test as was indicated in previous reports [23,26] we used the Pearson correlation coefficient and conducted bootstrapping (10,000 samples) to determine the 95% CI, thereby overcoming any possible confounding effect of the data’s distribution.

The velocity-variability relation was modeled for each of the step parameters using Linear modeling. For all step parameters, initial models included the terms “Group” (PwS and Healthy), “CWV,” and their interaction as predictors. Models also included the “CWV^2^” as an additional term. Non-significant terms were excluded from each model in a stepwise manner. Since the interaction term was non-significant in all the models, this term was excluded from the final models. For the step time, step length, and step width, parameter values were Log_e_ transformed to achieve normal distribution. For step velocity, we were unable to achieve normal distribution and therefore used a model based on Gamma distribution with a log link function. Finally, we tested the correlations of the residuals from the final models with the outcome measures of the LEMOCOT. To do so, we used linear modeling with the residuals as the predicted term, and “Group,” “LEMOCOT outcome,” and their interaction as predictors. In cases of a significant interaction term, we tested the correlation for each group independently. For statistical analysis we used the statistical package for social sciences (SPSS), version 26. Significance level was set a-priori to α<0.05.

## 3. Results

The participants’ characteristics are presented in Table 1. For 3 PwS the LEMOCOT score was 0. For these participants we were unable to calculate the absolute and variable error. One PwS performed the LEMOCOT with only one touch on the distal target, and therefore the variable error value was calculated for this participant based on the proximal target only.

**Table 1.** Participants’ characteristics

|  | **Controls**<br>**(n=20)** | **Stroke**<br>**(n=40)** | **$\Delta$** | **p-value** |
| --- | --- | --- | --- | --- |
| Sex *male/female* | 7/13 | 30/10 |  | 0.004 |
| Age *years* | 34 (6) | 65 (12) | -31 | <0.001 |
| Hight *cm* | 167.8 (2.4) | 167.4 (1.6) | 0.4 | 0.9 |
| Mass *kg* | 66.8 (3.2) | 74.6 (2.4) | -7.8 | 0.048 |
| Time after stroke onset *days* | ----- | 56 (6.5) | ----- | ----- |
| Preferred walking speed *m/sec* | 1.31 (0.24) | 0.72 (0.35) | 0.59 | <0.001 |
| Step time *SD msec* | 12 (4.5) | 65.1 (85.5) | -53.1 | <0.001 |
| Step length *SD mm* | 28.5 (45.3) | 47 (31.2) | -18.5 | <0.001 |
| Step width *SD mm* | 23.2 (4.6) | 19.2 (10.6) | 4 | 0.003 |
| Step velocity *SD mm/sec* | 60 (95) | 67 (46) | -7 | 0.005 |
| LEMOCOT score (*n*) | 38 (10) | 14 (9) | 24 | <0.001 |
| LEMOCOT absolute error *mm* | 11.2 (2.4) | 19.3 (13.1) | -8.1 | 0.002 |
| LEMOCOT variable error *mm* | 9.3 (1.7) | 13.5 (8) | -4.2 | 0.088 |

Values indicate mean (SD). Abbreviations: LEMOCOT, lower extremity motor coordination test.

### LEMOCOT and gait velocity

The correlations between the LEMOCOT outcome measures (i.e., score, absolute error, and variable error) and the CWV are presented in Table 2. Our results, consistent with previous reports [23,26], indicate that CWV and performance in the LEMOCOT correlate, but differences were observed between PwS and healthy adults.

**Table 2.** Correlations between preferred walking velocity and the LEMOCOT outcome measures

|  | **Healthy** |  | **Stroke** |  |
| --- | --- | --- | --- | --- |
|  | coefficient | 95% CI | coefficient | 95% CI |
| Score | -0.19 | -0.75-0.37 | 0.53 | 0.23-0.73 |
| Absolute error | -0.44 | -0.71-(-0.12) | -0.29 | -0.55-0.12 |
| Variable error | -0.49 | -0.74-(-0.16) | -0.18 | -0.46-0.17 |

### Gait velocity and spatiotemporal variability

The final velocity-variability models are presented in Table 3 and in Figure 1. Briefly, excluding step velocity SD, all parameters correlated with CWV. Specifically, for step time and step width a quadratic velocity-variability relation was observed while for step length a linear relation was observed (after the exclusion of a single highly influential datapoint from a healthy participant, i.e., Cook’s distance>1). After removal of a highly influential datapoint (from the same participant as above), the model for step velocity revealed no velocity-variability dependency. In all models, the ‘Group’ term was found significant after controlling for walking velocity, indicating that PwS exhibited greater variability in the step time, step length, and step velocity than did the healthy adults. For step width the opposite was observed (PwS exhibited less variability than healthy adults).

**Table 3.** Terms and coefficients of the STV by velocity models

|  | **Constant**<br>**(95%CI)** | **Group (95%CI)** | **Velocity (95%CI)** | **Velocity<sup>2</sup>**<br>**(95%CI)** | **p-value** |
| --- | --- | --- | --- | --- | --- |
| *Step time* | 5.2 (5.6-5.8) | 0.41 (0.08-0.75) | -3.6 [-4.8-(-2.5)] | 1.1 (0.5-1.8) | <0.001 |
| *Step length* | 3.9 (3.3-4.4) | 3.5 (0.01-0.69) | -0.8 [-1.2-(-0.4)] |  | <0.001 |
| *Step width* | 4 (3.5-4.5) | -0.41 [-0.68-(-0.14)] | -1.8 [-2.8-(-0.9)] | 0.86 (0.3-1.4) | <0.001 |
| *Step velocity* | -3.2 [-3.5-(-3)] | 0.55 (0.28-0.83) |  |  | <0.001 |

**Figure 1.**
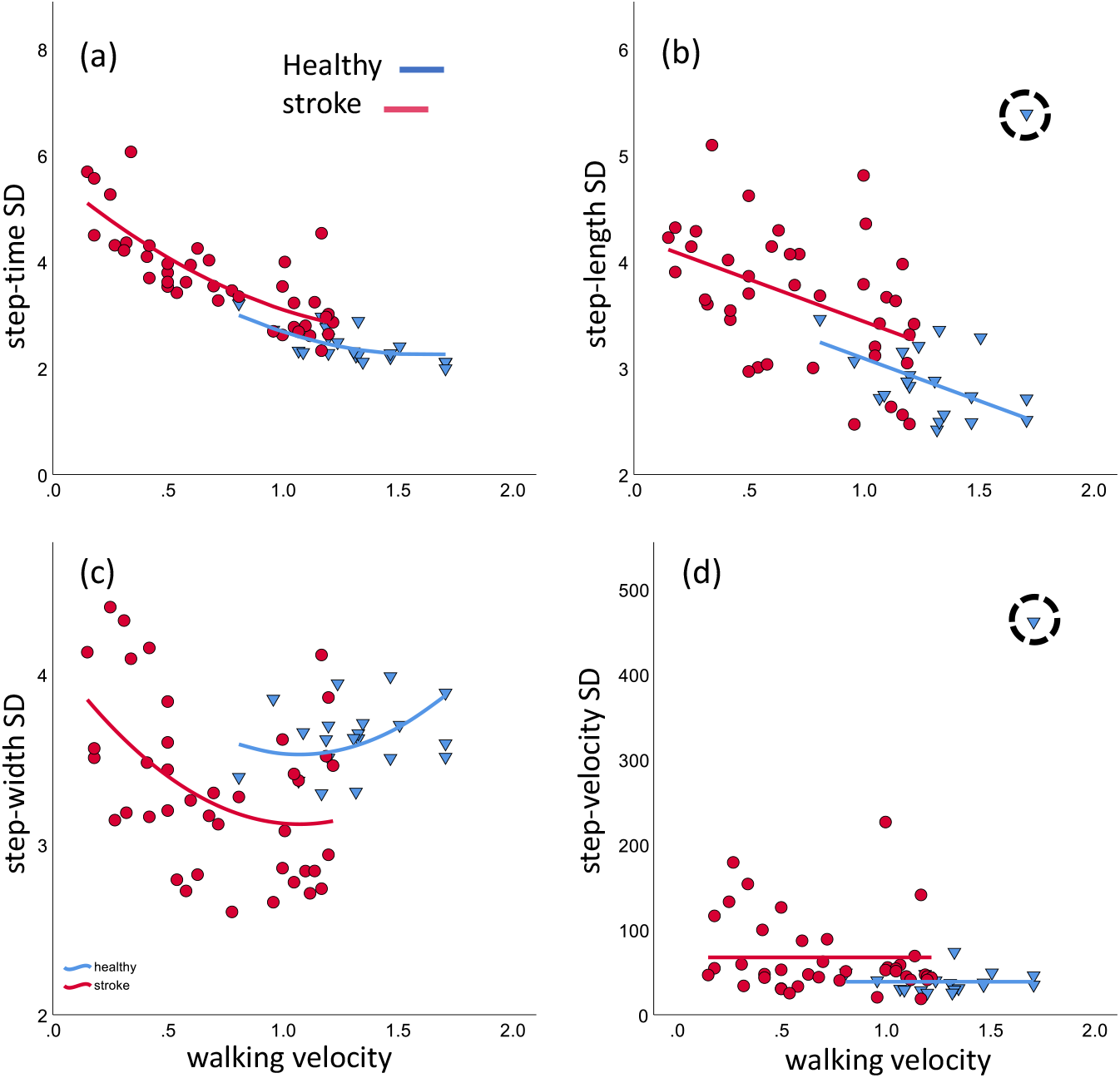
The fitted models—red and blue lines for persons with stroke (PwS) and healthy adults, respectively—of the gait velocity-variability relation for each step parameter: step-time variability (left upper), step-length variability (right upper), step-width variability (lower left), and step-velocity variability (lower right). The Y-axis values are the log_e_ transformation of the actual values in all panels except for step velocity for which Y-axis values are presented in mm/sec. X-axis values in all panels are presented in m/sec. Red circles and blue triangles represent data of PwS and healthy adults, respectively. The dashed-line circle in the right panels highlights the extreme data point that was excluded from the models.

### Spatiotemporal variability and LEMOCOT

The results of the significant models are depicted in Figure 2. The analysis revealed that after controlling for walking velocity, the LEMOCOT outcome measures correlated with the step parameters, but only in PwS. Specifically, the LEMOCOT score correlated with step time variability in PwS (β=-0.022, 95%CI [-0.04-(-0.003)], p=0.02) but not in healthy adults (p=0.72), indicating that in PwS poor performance on the LEMOCOT was associated with increased step-time variability. None of the other STV parameters correlated with the number of touches.

**Figure 2.**
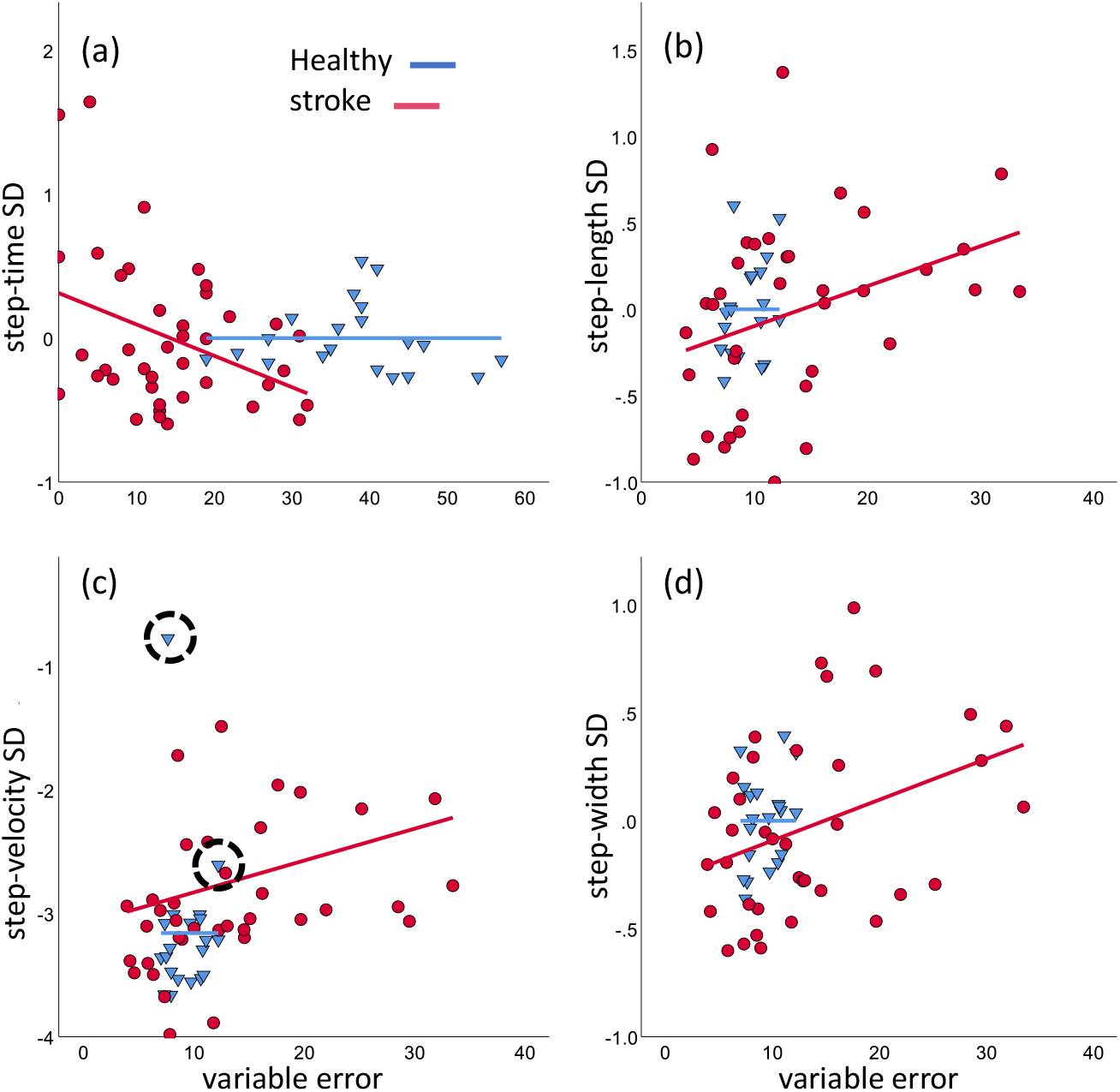
The correlation between the LEMOCOT outcome measures and step-variability parameters after controlling for gait velocity in healthy adults (blue circles) and persons with stroke (PwS) (red triangles). Red and blue lines for PwS and healthy adults respectively, represent the fitted models for (a) step-time variability by LEMOCOT score, (b) step-length variability by LEMOCOT variable error, (c) step-velocity variability by LEMOCOT variable error, and (d) step-width variability by LEMOCOT variable error. The values in panel c are presented as log_e_ transformations, as the Gamma regression uses a log_e_ link function. The dashed-line circle in panel c highlights the extreme data points that were excluded from the models.

For the absolute error in the LEMOCOT we found no significant correlation with any of the velocity-controlled step parameters. For the variable error in the LEMOCOT we found a significant correlation with the velocity-controlled step-width variability (β=0.02, 95%CI [0.002-0.036], p=0.03) and step-length variability (β=0.023, 95%CI [0.001-0.045], p=0.04) in PwS but not in healthy adults. A significant correlation between the variable error in the LEMOCOT and step velocity variability was found in PwS (β=0.026, 95%CI [0.003-0.049], p=0.03), but not in healthy adults (after removing two highly influential data-points).

## 4. Discussion

In this study, we explored whether greater STV during walking is directly related to deficits in motor coordination of the paretic lower limb in PwS. The main findings from this investigation were that STV differed between PwS and healthy adults regardless of their walking velocity and that the STV is directly related to the ability of PwS to coordinate the movements of their affected lower extremity.

Previous reports have indicated that PwS walk at a slower velocity and with greater STV than do healthy individuals [17,18]. Decreased walking velocity and increased STV are, independently, considered indicative of poor motor control and instability, and serve as a biomarker for pathology and as a predictor for future falls. Nevertheless, it is well documented that STV is sensitive to walking velocity [11-14], making it unclear whether the increased STV observed in PwS simply reflects their slow walking velocity. Also, it is unclear whether increased STV is a general biomarker for pathology or a quantitative measure of motor control deficits.

A recent report [28] attempted to separate the effect of stroke on STV from that of walking velocity, and the results agreed with ours in part. Specifically, these authors found that only step-time variability differed between PwS and healthy controls, after adjusting for walking velocity. We believe that the difference between our results and theirs is due to methodological differences. Mainly, these authors tested overground walking and used the coefficient of variation as their variability measure. Further, they controlled for velocity by using both participant matching and by adding velocity as a covariate in their model, but as a linear term. Our data clearly indicate that the velocity-variability relation of all gait parameters is exponential, suggesting that estimates produced using their methods are likely inaccurate. For future investigation, we provide here the full models used to estimate STV from velocity including the CI of the coefficients.

Regarding our main objective, we found that the performance in the LEMOCOT correlated with all STV parameters after controlling for velocity, but only in PwS. The direction of this correlation consistently related better performance in the LEMOCOT with reduced gait STV. This indicates that STV is not merely a general biomarker of pathology but is instead sensitive to the level of coordination deficit. Interestingly, we found that the LEMOCOT score correlated with temporal variability (step time) but not with spatial variability (step length and step width). This was an unexpected finding as the task requires both temporal and spatial accuracy. In contrast, the variable error in the LEMOCOT that quantifies spatial accuracy was correlated with the spatial parameters of gait variability (step-length variability and step-width variability) but not with temporal variability. Further, variability in step velocity, which captures both temporal and spatial domains, was correlated only with accuracy. Handelzalts et al. [27] suggested that in the LEMOCOT examiners count leg movements and not “in-target touches.” If true, then such measure emphasizes the performance in the temporal domain neglecting its spatial domain. The authors suggested additional outcome measures that emphasize the accuracy in the spatial domain [27]. The results of the current report do suggest that this addition is important (at least in terms of precision, as reflected by the VE), as these measures quantify all movements preformed (in- and out-of-target touches) as opposed to a binary classifying of the traditional score.

Recently, Patel et al. [29] reported that both temporal and spatial gait variability correlated with inter-limb coordination of walking in PwS, signifying that not only intra-limb coordination is required for safe and stable walking. The observed relations in both reports should be further examined to test whether these relations are causal, because if this is the case, rehabilitation programs for PwS would benefit from adding intra- and inter-limb coordination training.

Finally, although this is not the main objective of the current investigation, a significant velocity-variability correlation was found for all step parameters, except for step velocity. The reason for this relation remains an open question in research. One may speculate that both walking velocity and gait STV capture independent aspects of motor control. In other words, this correlation is not causal but merely relational. This possibility is unlikely since the within-subject effect of walking velocity has been shown to have a great impact on STV in both healthy and pathological populations [11-16] indicating that this relation is, at least in part, causal. Another possibility is that STV exclusively represents executional errors. If so, the control system might allow some degree of error that is proportional to the magnitude of the desired outcome. In fact, according to Weber’s law, the ability to sense change in a signal is proportional to its magnitude, meaning that the control system does not “allow” errors, but is simply unable to perceive and subsequently correct them. This explanation, however, has several problems. First, the true nature of STV seems to be much more complicated than simply errors in execution [30]. That is, variability is likely to represent both errors and flexibility, meaning that some level of variability is actually desirable [8,30]. Second, while steps shorten temporally as velocity increases, they become spatially longer, but both temporal and spatial variability decreased as walking velocity increased (see Figure 1 a,b). Another possible explanation for this relation is that the control system does not try to actively control these variables, but rather their fluctuations are dictated by the forces acting on the system. In that case, as gait velocity increases, so does the system’s momentum, making it more stable (i.e., more energy is required to change its state). This possibility has some support from the uncontrolled manifold theory [31,32]. According to this theory, state variables can fluctuate, and will not be actively controlled as long as these fluctuations do not affect the desired value of the state variable of interest. While what this/these state-variable/s might be in walking is largely unknow, in the special case of treadmill walking it might be the step velocity [33,34]. This may explain why variability in step velocity was unrelated to the CWV in the current investigation. While there might be other explanations as well as combinations of the above explanations, overall, the true nature of this variability-velocity relation is not well understood.

## 5. Limitations

This investigation has several limitations. One important limitation is the fact that all PwS walked while holding the handrails while none of the healthy adults did. Holding the handrails provides greater support and stability that might have affected the magnitude of the STV observed, especially for step width. While current literature does support our finding of reduced variability in step width [8], it is possible that this observation reflects the greater support provided by the handrails. Another limitation is that our control group was not age matched to the PwS group. Due to this age difference, we cannot conclude with certainty that the difference between groups was derived exclusively from deficits caused by brain damage.

## 6. Conclusions

We found that decreased performance in the LEMOCOT was associated with increased STV in PwS, even after controlling for walking velocity. Our results demonstrate the connection between lower-extremity intra-limb coordination impairments and deficits in gait function.

## Data Availability

All data produced in the present study are available upon reasonable request to the authors

## References

[1] Batchelor FA, Mackintosh SF, Said CM, Hill KD. Falls after Stroke. International Journal of Stroke 2012;76:482–90.

[2] Weerdesteyn V, de Niet M, van Duijnhoven HJR, Geurts ACH. Falls in individuals with stroke. J Rehabil Res Dev 2008;458:1195–213.

[3] Bruijn SM, Meijer OG, Beek PJ, van Dieen JH. Assessing the stability of human locomotion: a review of current measures. J R Soc Interface 2013;1083:20120999.

[4] Hausdorff JM. Gait dynamics, fractals and falls: finding meaning in the stride-to-stride fluctuations of human walking. Hum Mov Sci 2007;264:555–89.

[5] Hamacher D, Singh NB, Van Dieën JH, Heller MO, Taylor WR. Kinematic measures for assessing gait stability in elderly individuals: a systematic review. Journal of The Royal Society Interface 2011;865:1682–98.

[6] Owings TM, Grabiner MD. Variability of step kinematics in young and older adults. Gait Posture 2004;20.

[7] Konig N, Taylor WR, Baumann CR, Wenderoth N, Singh NB. Revealing the quality of movement: A meta-analysis review to quantify the thresholds to pathological variability during standing and walking. Neurosci Biobehav Rev 2016;68:111–9.

[8] Ravi DK, et al. Revealing the optimal thresholds for movement performance: A systematic review and meta-analysis to benchmark pathological walking behaviour. Neurosci Biobehav Rev 2020;108:24–33.

[9] Maki BE. Gait changes in older adults: predictors of falls or indicators of fear. J Am Geriatr Soc 1997;453:313–20.

[10] Hausdorff JM, Rios DA, Eldelberg HK. Gait variability and fall risk in community-living older adults: a 1-year prospective study. Arch Phys Med Rehabil 2001;82.

[11] Jordan K, Challis JH, Newell KM. Walking speed influences on gait cycle variability. Gait Posture 2007;26.

[12] Kang HG, Dingwell JB. Effects of walking speed, strength and range of motion on gait stability in healthy older adults. J Biomech 2008;4114:2899–905.

[13] Beauchet O, et al. Walking speed-related changes in stride time variability: effects of decreased speed. J Neuroeng Rehabil 2009;6:32-.

[14] Chien JH, Yentes J, Stergiou N, Siu KC. The Effect of Walking Speed on Gait Variability in Healthy Young, Middle-aged and Elderly Individuals. J Phys Act Nutr Rehabil 2015;2015 Epub 2015;2015.

[15] Frenkel-Toledo S, Giladi N, Peretz C, Herman T, Gruendlinger L, Hausdorff JM. Effect of gait speed on gait rhythmicity in Parkinson’s disease: variability of stride time and swing time respond differently. J Neuroengineering Rehabil 2005;2.

[16] Kao PC, Dingwell JB, Higginson JS, Binder-Macleod S. PMC4251664; Dynamic instability during post-stroke hemiparetic walking. Gait Posture 2014;40; 2014/06/173:457-63.

[17] Balasubramanian CK, Neptune RR, Kautz SA. PMC2675553; Variability in spatiotemporal step characteristics and its relationship to walking performance post-stroke. Gait Posture 2009;29; 2008/12/063:408-14.

[18] Wang Y, et al. Gait characteristics of post-stroke hemiparetic patients with different walking speeds. Int J Rehabil Res 2020;431:69–75.

[19] Tomita Y, Rodrigues MRM, Levin MF. Upper Limb Coordination in Individuals With Stroke: Poorly Defined and Poorly Quantified. Neurorehabil Neural Repair 2017;3110-11:885–97.

[20] Alouche SR, Molad R, Demers M, Levin MF. Development of a Comprehensive Outcome Measure for Motor Coordination; Step 1: Three-Phase Content Validity Process. Neurorehabil Neural Repair 2021;352:185–93.

[21] Srivastava S, Kao P, Reisman DS, Higginson JS, Scholz JP. Coordination of muscles to control the footpath during over-ground walking in neurologically intact individuals and stroke survivors. Exp Brain Res 2016;2347:1903–14.

[22] Lofrumento M, et al. Effects of gait rehabilitation on motor coordination in stroke survivors: an UCM-based approach. Exp Brain Res 2021;2397:2107–18.

[23] Desrosiers J, Rochette A, Corriveau H. Validation of a new lower-extremity motor coordination test. Arch Phys Med Rehabil 2005;86; 2005/05/175:993-8.

[24] Hackett ML, Duncan JR, Anderson CS, Broad JB, Bonita R. Health-related quality of life among long-term survivors of stroke : results from the Auckland Stroke Study, 1991-1992. Stroke 2000;312:440–7.

[25] Latash ML, Anson JG. Synergies in health and disease: relations to adaptive changes in motor coordination. Phys Ther 2006;868:1151–60.

[26] Kwan MS, Hassett LM, Ada L, Canning CG. PMC6849089; Relationship between lower limb coordination and walking speed after stroke: an observational study. Braz J Phys Ther 2019;23; 2019/11/126:527-31.

[27] Handelzalts S, et al. Insights into motor performance deficits after stroke: an automated and refined analysis of the lower-extremity motor coordination test (LEMOCOT). Journal of neuroengineering and rehabilitation 2021;181:1–10.

[28] Chow JW, Stokic DS. The contribution of walking speed versus recent stroke to temporospatial gait variability. Gait Posture 2022;100:216–21.

[29] Patel P, Enzastiga D, Casamento-Moran A, Christou EA, Lodha N. Increased temporal stride variability contributes to impaired gait coordination after stroke. Sci Rep 2022;121:12679.

[30] Cavanaugh JT, Stergiou N. Gait variability: a theoretical framework for gait analysis and biomechanics. Biomechanics and Gait Analysis 2020;274:251.

[31] Scholz JP, Schöner G. The uncontrolled manifold concept: identifying control variables for a functional task. Experimental brain research 1999;1263:289–306.

[32] Latash ML, Scholz JP, Schöner G. Motor control strategies revealed in the structure of motor variability. Exerc Sport Sci Rev 2002;301:26–31.

[33] Dingwell JB, Cusumano JP. Identifying stride-to-stride control strategies in human treadmill walking. PloS one 2015;104:e0124879.

[34] Dingwell JB, John J, Cusumano JP. Do humans optimally exploit redundancy to control step variability in walking? PLoS Comput Biol 2010;67:e1000856.

